# COVID Outcome Prediction in the Emergency Department (COPE): Development and validation of a model for predicting death and need for intensive care in COVID-19 patients

**DOI:** 10.1101/2020.12.30.20249023

**Authors:** David van Klaveren, Alexandros Rekkas, Jelmer Alsma, Rob JCG Verdonschot, Dick TJJ Koning, Marlijn JA Kamps, Tom Dormans, Robert Stassen, Sebastiaan Weijer, Klaas-Sierk Arnold, Benjamin Tomlow, Hilde RH de Geus, Rozemarijn L van Bruchem-Visser, Jelle R Miedema, Annelies Verbon, Els van Nood, David M Kent, Stephanie CE Schuit, Hester Lingsma

**Affiliations:** Department of Public Health, Erasmus University Medical Center Rotterdam, The Netherlands; Predictive Analytics and Comparative Effectiveness Center, Institute for Clinical Research and Health Policy Studies, Tufts Medical Center, Boston, USA; Department of Medical Informatics, Erasmus University Medical Center Rotterdam, The Netherlands; Department of Internal Medicine, Erasmus University Medical Center, Rotterdam, The Netherlands; Emergency Department, Erasmus University Medical Center, Rotterdam, The Netherlands; Department of Intensive Care, Catharina Hospital, Eindhoven, The Netherlands; Department of Intensive Care, Zuyderland Medical Center, Heerlen, Limburg, The Netherlands; Department of Traumatology, Maastricht University Medical Centre, The Netherlands; Department of Internal Medicine, Antonius Hospital, Sneek, The Netherlands; Department of Intensive Care, Antonius Hospital, Sneek, The Netherlands; Department of Pulmonary Medicine, Isala Clinics, Zwolle, The Netherlands; Department of Intensive Care, Erasmus University Medical Center, Rotterdam, The Netherlands; Department of Pulmonary Medicine, Erasmus University Medical Center, Rotterdam, The Netherlands; Department of Medical Microbiology and Infectious Diseases, Erasmus University Medical Center, Rotterdam, The Netherlands; University Medical Center Groningen, The Netherlands

**Author notes:** **Corresponding author:** David van Klaveren, Department of Public Health, Erasmus University Medical Center Rotterdam, Dr. Molewaterplein 50, 3015 GE ROTTERDAM, The Netherlands.

**Keywords:** COVID-19, Emergency Department, Clinical Prediction Models

## Abstract

**Background and aim:** COVID-19 is putting extraordinary pressure on emergency departments (EDs). To support decision making in the ED, we aimed to develop a simple and valid model for predicting mortality and need for intensive care unit (ICU) admission in suspected COVID-19 patients.

**Methods:** For model development, we retrospectively collected data of patients who were admitted to 4 large Dutch hospitals with suspected COVID-19 between March and August 2020 (first wave of the pandemic). Based on prior literature we considered quickly and objectively obtainable patient characteristics, vital parameters and blood test values as predictors. Logistic regression analyses with post-hoc uniform shrinkage was used to obtain predicted probabilities of in-hospital death and of the need for ICU admission, both within 28 days after hospital admission. We assessed model performance (Area Under the ROC curve (AUC); calibration plots) with temporal validation in patients who presented between September and December 2020 (second wave). We used multiple imputation to account for missing values.

**Results:** The development data included 5,831 patients, of whom 629 (10.8%) died and 5,070 (86.9%) were discharged within 28 days after admission. ICU admission was fully recorded for 2,633 first wave patients in 2 hospitals, with 214 (8%) ICU admissions within 28 days. A simple model – COVID Outcome Prediction in the Emergency Department (COPE) – with age, respiratory rate, C-reactive protein, lactic dehydrogenase, albumin and urea captured most of the ability to predict death. COPE was well-calibrated and showed good discrimination in 3,252 second wave patients (AUC in 4 hospitals: 0.82 [0.78; 0.86]; 0.82 [0.74; 0.90]; 0.79 [0.70; 0.88]; 0.83 [0.79; 0.86]). COPE was also able to identify patients at high risk of needing IC in 706 second wave patients with complete information on ICU admission (AUC: 0.84 [0.78; 0.90]; 0.81 [0.66; 0.95]). The models are implemented in web-based and mobile applications.

**Conclusion:** COPE is a simple tool that is well able to predict mortality and need for ICU admission for patients who present to the ED with suspected COVID-19 and may help patients and doctors in decision making.

**CONTRIBUTION TO THE LITERATURE:** *What is already known on this topic:*

- Prediction models have the potential to support decision making about hospital admission of patients presenting to the emergency department with suspected COVID-19
- Most currently available models that were independently assessed contain a high risk of bias
- Promising models were developed in different patient selections and included predictors that are not quickly and objective obtainable in emergency departments

*What this study adds:*

- A simple and objective tool (“COPE”) is well able to predict mortality and need for ICU admission for patients who present to the ED with suspected COVID-19
- COPE may support ED physicians to identify high-risk patients – i.e. those at high risk of deterioration and/or death – requiring treatment in the ICU, intermediate-risk patients requiring admission to the clinical ward, and low-risk patients who can potentially be sent home

## BACKGROUND

The COVID-19 pandemic is putting extraordinary pressure on emergency departments (EDs), clinical wards and intensive care units (ICUs). Clinical prediction models for COVID-19 outcomes have the potential to support decision making about hospital admission. But most currently available models that were assessed with the prediction model risk of bias assessment tool (PROBAST) contain a high risk of bias (1-3). The most common reasons were non-representative selection of control patients, exclusion of patients in whom the event of interest was not observed by the end of the study, high risk of model overfitting, and vague reporting. Additionally, the description of the study population or intended use of the models was often missing, and calibration of the model predictions was rarely assessed.

The recently proposed 4C Mortality Score is probably at low risk of bias, but was derived from a selected population of patients admitted to UK hospitals who were seriously ill (mortality rate of 32.2%). Predictors included the number of comorbidities and the Glasgow Coma Scale, items that are not easily and unambiguously obtained for patients with suspected COVID-19 at EDs everywhere (4, 5). Similarly, the promising risk scores VACO and COVID-GRAM – predicting 30-day mortality in positively tested patients and critical illness in hospitalized patients, respectively – require knowledge on pre-existing comorbidities (6, 7). The COVID-GRAM model also requires chest radiography results.

We aimed to develop and validate a simple and valid model for predicting mortality and the need for ICU in all patients who are suspected to have COVID-19 when presenting at the ED. To facilitate implementation in clinical practice, we only included quickly and objectively obtainable patient characteristics, vital parameters and blood test values.

## METHODS

### Population

19 large Dutch hospitals were requested to supply retrospective data on the cohorts of COVID-19 patients who were admitted to their hospital. Of those hospitals, Catharina Hospital Eindhoven, Zuyderland Medical Center Heerlen, Isala Clinics Zwolle, Erasmus University Medical Center Rotterdam and Antonius Hospital Sneek supplied these data. The data from Antonius Hospital Sneek were not used in the analyses, because of large proportions of missing predictor values.

For model development, we used the data of patients who presented at the ED and were admitted to the hospital with suspected COVID-19 in the first wave of the pandemic, that is from March up to and including August 2020. Patients being transferred to other hospitals were excluded since information on outcomes was missing. For model validation we used data of patients who presented at the ED and were admitted to the hospital with suspected COVID-19 in the second wave of the pandemic, that is from September up to and including December 2020. We used Multivariate Imputation by Chained Equations (R-packages mice) for multiple imputation of missing predictor values (8, 9).

### Outcomes

The outcomes of interest were in-hospital death and admission to ICU within 28 days after hospital admission. Transfer to a hospice was counted as death.

### Predictors

Based on prior literature we included patient characteristics (sex, age, BMI), vital parameters (oxygen saturation, systolic blood pressure, heart rate, respiratory rate [RR], body temperature) and blood test values (C-reactive protein [CRP], lactic dehydrogenase [LDH], D-Dimer, leucocytes, lymphocytes, monocytes, neutrophils, eosinophils, MCV, albumin, bicarbonate, sodium, creatinine,, urea), all measured at ED admission, as potential predictors (1). Furthermore, we included the month of admission to capture potential changes in outcomes over time.

### Model development

Logistic regression was used to analyze associations between predictors and outcomes. We decided on including non-linear transformations of potential predictors on the basis of a full model with a restricted cubic spline (3 knots; 2 regression coefficients) for each continuous predictor (10, 11). Based on Wald statistics, we selected the most promising predictors into a parsimonious model for easy use in clinical practice. To prevent overfitting, we used bootstrap validation – including the same variable selection strategy to mimic our modeling strategy – to estimate a uniform shrinkage factor (11). The regression coefficients of the final model were multiplied by this shrinkage factor, and the model intercept was adjusted to ensure overall calibration of the model. We used the R-package rms (Regression Modeling Strategies) for regression analyses (8, 12).

### Model validation

Model performance was assessed with temporal validation in second wave patients, in each of the 4 separate hospitals. We assessed discriminative ability with the area under the operator receiver characteristic curve (AUC) and calibration with calibration plots of five equally sized groups of predicted risk, calibration intercepts, and calibration slopes. The model-based concordance (mbc) was used to understand the impact of potential differences in case-mix heterogeneity between the development and validation data on discriminative ability (13).

### Patient and Public Involvement

Patients were not directly involved in the design of this study. The outcome of interest and the potential predictors were selected up front by a group of hospital physicians caring for COVID-19 patients (ED, internal medicine, pulmonary medicine, ICU). Since we retrospectively collected data, patients were not burdened by our study. In future research, we will convene multi-stakeholder panels of approximately 12 members including COVID-19 patients, relatives, hospitals physicians caring for COVID-19 patients, palliative care physicians, and ethicists, with the aim to develop a full understanding of how the models may best support patients and clinicians in making critical patient-centered decisions.

## RESULTS

### Population and outcomes

The database contained 5,912 patients who presented at the ED from March up to and including August 2020 and who were admitted to the hospital with a suspicion of COVID-19. Of those patients 81 (1.4%) were excluded because of a transfer to other hospitals (outcome not recorded). The development data included 5,831 patients of whom 629 (10.8%) died, 5,070 (86.9%) were discharged within 28 days after hospital admission, and 132 (2.3%) were still in hospital at 28 days after admission. Patients who died – in comparison with patients who were discharged – tended to be more often male, at older age, with aberrant vital parameters (higher RR and HR; lower oxygen saturation), higher blood levels of CRP, LDH, creatinine and urea and lower blood levels of lymphocytes and albumin (Table 1).

**Table 1.** Baseline characteristics of development and validation patient cohorts. Median (“M”) and quartile range (“Q1” = first quartile; “Q3” = third quartile) are presented for all continuous variables. Status, “All patients”, “Discharged”, “In hospital” and “Dead” is measured at 28 days after hospital admission.

|  | % missing | All patients |  |  | Discharged |  |  | In hospital |  |  | Dead |  |  |
| --- | --- | --- | --- | --- | --- | --- | --- | --- | --- | --- | --- | --- | --- |
| **A. Development data** |  | n = 5831 |  |  | n = 5070 |  |  | n = 132 |  |  | n = 629 |  |  |
| Male sex | 0 | 57 | % |  | 56 | % |  | 64 | % |  | 64 | % |  |
|  |  | M | Q1 | Q3 | M | Q1 | Q3 | M | Q1 | Q3 | M | Q1 | Q3 |
| Age (years) | 0 | 70 | 58 | 80 | 69 | 56 | 78 | 71 | 62 | 79 | 78 | 70 | 84 |
| BMI (kg/m<sup>2</sup>) | 58 | 26 | 23 | 30 | 26 | 23 | 30 | 25 | 23 | 29 | 26 | 23 | 30 |
| HR (bpm) | 39 | 90 | 78 | 103 | 90 | 78 | 103 | 87 | 76 | 99 | 93 | 80 | 107 |
| SBP (mmHg) | 42 | 133 | 118 | 150 | 133 | 119 | 151 | 136 | 115 | 152 | 131 | 114 | 145 |
| RR (/min) | 42 | 19 | 16 | 23 | 19 | 16 | 23 | 20 | 17 | 24 | 23 | 19 | 28 |
| Saturation (%) | 41 | 95.8 | 94.0 | 97.5 | 96.0 | 94.3 | 97.8 | 95.4 | 93.6 | 97.0 | 94.1 | 91.9 | 96.0 |
| Temperature (°C) | 40 | 37.3 | 36.7 | 38.1 | 37.3 | 36.7 | 38.0 | 37.5 | 37.0 | 38.1 | 37.4 | 36.7 | 38.1 |
| CRP (mg/L) | 7 | 48 | 10 | 118 | 43 | 8 | 110 | 85 | 34 | 160 | 91 | 41 | 180 |
| D dimer (µg/L) | 64 | 1100 | 527 | 2545 | 1028 | 504 | 2300 | 1950 | 719 | 8110 | 2100 | 949 | 4772 |
| LDH (U/L) | 18 | 244 | 200 | 322 | 237 | 197 | 302 | 300 | 234 | 422 | 338 | 253 | 492 |
| Leucocytes (x10<sup>9</sup>/L) | 7 | 9.1 | 6.7 | 12.7 | 9.1 | 6.7 | 12.6 | 9.4 | 6.5 | 12.5 | 9.2 | 6.4 | 14.0 |
| Lymphocytes | 16 | 1.04 | 0.66 | 1.6 | 1.10 | 0.7 | 1.7 | 0.80 | 0.54 | 1.3 | 0.80 | 0.51 | 1.3 |
| Albumin (g/L) | 15 | 39 | 35.5 | 42 | 39.8 | 36 | 42.8 | 37 | 34 | 41 | 36 | 33 | 39 |
| Bicarbonate (mmol/L) | 45 | 23.6 | 21 | 26 | 23.8 | 22 | 26 | 22.8 | 21 | 26 | 22.3 | 20 | 25 |
| Creatinine (µmol/L) | 8 | 84 | 66 | 111 | 82 | 65 | 107 | 89 | 66 | 111 | 102 | 75 | 153 |
| Eosinophils (x10<sup>9</sup>/L) | 26 | 0.03 | 0.00 | 0.10 | 0.03 | 0.01 | 0.10 | 0.03 | 0.00 | 0.10 | 0.01 | 0.00 | 0.10 |
| MCV (fL) | 7 | 90 | 87 | 94 | 90 | 86 | 94 | 90 | 88 | 94 | 91 | 87 | 96 |
| Monocytes (x10<sup>9</sup>/L) | 30 | 0.67 | 0.44 | 0.95 | 0.68 | 0.45 | 0.95 | 0.59 | 0.33 | 0.83 | 0.61 | 0.33 | 0.94 |
| Neutrophils (x10<sup>9</sup>/L) | 16 | 5.6 | 2.2 | 9.0 | 5.6 | 2.2 | 8.9 | 7.0 | 4.0 | 10.3 | 5.5 | 1.8 | 9.1 |
| Sodium (mmol/L) | 9 | 138 | 135 | 140 | 138 | 135 | 140 | 137 | 133 | 141 | 137 | 134 | 140 |
| Urea (mmol/L) | 9 | 6.5 | 4.6 | 9.7 | 6.2 | 4.5 | 9.0 | 7.4 | 5.1 | 11.0 | 9.6 | 6.6 | 15.0 |
| **B. Validation data** |  | n = 3252 |  |  | n = 2854 |  |  | n = 72 |  |  | n = 326 |  |  |
| Male sex | 0 | 56 | % |  | 56 | % |  | 46 | % |  | 61 | % |  |
|  |  | M | Q1 | Q3 | M | Q1 | Q3 | M | Q1 | Q3 | M | Q1 | Q3 |
| Age (years) | 0 | 71 | 58 | 80 | 69 | 55 | 79 | 72 | 58 | 82 | 79 | 73 | 85 |
| BMI (kg/m<sup>2</sup>) | 59 | 26 | 23 | 30 | 26 | 23 | 30 | 26 | 24 | 30 | 25 | 22 | 29 |
| HR (bpm) | 40 | 90 | 78 | 105 | 90 | 78 | 104 | 84 | 75 | 106 | 92 | 78 | 105 |
| SBP (mmHg) | 43 | 134 | 119 | 151 | 135 | 120 | 152 | 134 | 122 | 149 | 129 | 110 | 141 |
| RR (/min) | 43 | 20 | 16 | 24 | 20 | 16 | 24 | 20 | 17 | 26 | 23 | 19 | 27 |
| Saturation (%) | 40 | 95.7 | 94.0 | 97.5 | 96.0 | 94.0 | 97.7 | 95.5 | 94.0 | 97.0 | 94.8 | 92.3 | 96.5 |
| Temperature (°C) | 42 | 37.3 | 36.7 | 38.1 | 37.3 | 36.7 | 38.1 | 37.3 | 36.8 | 38.1 | 37.2 | 36.4 | 38.0 |
| CRP (mg/L) | 9 | 57 | 16 | 124 | 54 | 15 | 120 | 76 | 21 | 169 | 80 | 33 | 159 |
| D dimer (µg/L) | 76 | 1060 | 531 | 2170 | 1013 | 490 | 2012 | 1080 | 640 | 2570 | 1495 | 870 | 3724 |
| LDH (U/L) | 22 | 247 | 203 | 334 | 242 | 200 | 317 | 281 | 226 | 390 | 315 | 238 | 489 |
| Leucocytes (x10<sup>9</sup>/L) | 10 | 9.4 | 6.6 | 12.9 | 9.4 | 6.6 | 12.9 | 9.6 | 7.0 | 13.2 | 9.5 | 6.8 | 13.3 |
| Lymphocytes | 20 | 0.98 | 0.62 | 1.50 | 1.00 | 0.64 | 1.50 | 1.10 | 0.72 | 1.53 | 0.71 | 0.48 | 1.20 |
| Albumin (g/L) | 20 | 38.7 | 34.9 | 41.8 | 39 | 35 | 42 | 35.95 | 32 | 38.9 | 36.2 | 31.9 | 40 |
| Bicarbonate (mmol/L) | 50 | 23.5 | 21 | 26 | 23.6 | 21 | 26 | 22.8 | 20 | 27 | 22.8 | 20 | 25 |
| Creatinine (µmol/L) | 10 | 84 | 66 | 116 | 83 | 65 | 111 | 80 | 58 | 118 | 103 | 74 | 158 |
| Eosinophils (x10<sup>9</sup>/L) | 27 | 0.03 | 0.01 | 0.10 | 0.03 | 0.01 | 0.10 | 0.03 | 0.01 | 0.09 | 0.03 | 0.00 | 0.04 |
| MCV (fL) | 10 | 90 | 87 | 94 | 90 | 87 | 94 | 92 | 88 | 95 | 91 | 88 | 96 |
| Monocytes (x10<sup>9</sup>/L) | 30 | 0.67 | 0.43 | 0.98 | 0.67 | 0.44 | 0.98 | 0.57 | 0.38 | 1.00 | 0.58 | 0.36 | 0.97 |
| Neutrophils (x10<sup>9</sup>/L) | 21 | 5.8 | 2.4 | 9.4 | 5.8 | 2.4 | 9.4 | 6.6 | 3.3 | 9.1 | 5.6 | 2.2 | 9.4 |
| Sodium (mmol/L) | 11 | 137 | 134 | 139 | 137 | 134 | 139 | 136 | 133 | 139 | 137 | 133 | 140 |
| Urea (mmol/L) | 11 | 6.9 | 4.9 | 10.4 | 6.6 | 4.7 | 9.8 | 7.3 | 5.3 | 11.8 | 10.3 | 7.4 | 17.0 |

Similar patterns were seen in 3.252 patients who were admitted to hospital in the second wave of the pandemic from September up to and including December 2020, of whom 326 (10.0%) died, 2,854 (87.8%) were discharged within 28 days after admission, and 72 (2.2%) were still in hospital at 28 days after admission. Admission to ICU was fully recorded – including ICU admissions at a later time point than the initial hospital admission – for 2,633 patients in 2 hospitals (214 ICU admissions within 28 days [8.1%]) in the first wave and in 1,466 patients (86 ICU admissions within 28 days [5.9%]) in the second wave of the pandemic.

### Prediction of death

Patients who were admitted in the first month of the pandemic in the Netherlands, that is in March 2020, were at substantially increased risk of death (Table 2: multivariable odds ratio 1.99; 95% confidence interval 1.61-2.47). All models included this correction factor for the first month, to avoid overestimation of risk after the first month of the pandemic.

**Table 2.** Univariable and multivariable associations between predictors and death within 28 days. Odds ratios (OR) with 95% confidence intervals (CI) for separate variables (columns “Univariable”), for a model with all available predictors (columns “Full model”) and for a model with only the six strongest predictors (columns “Selected model”). Variable importance is expressed with the Wald statistic (columns “Wald”). The odds ratios of the final model are based on the Selected model, with a uniform shrinkage factor of 0.93 (column “COPE”). Associations are based on 5,831 patients of whom 629 died within 28 days.

| Predictor | Contrast | Univariable |  |  |  | Full model |  |  |  | Selected model |  |  |  | COPE |
| --- | --- | --- | --- | --- | --- | --- | --- | --- | --- | --- | --- | --- | --- | --- |
|  |  | OR | 95% CI |  | Wald | OR | 95% CI |  | Wald | OR | 95% CI |  | Wald | OR |
| Month | ≥April vs <April | 2.57 | 2.16 | 3.05 | 114 | 1.99 | 1.61 | 2.47 | 39 | 2.06 | 1.68 | 2.52 | 49 | 1.96 |
| Sex | male vs female | 1.37 | 1.15 | 1.63 | 13 | 1.12 | 0.90 | 1.39 | 1 |  |  |  |  |  |
| Age (years) | 80 vs 58 | 3.07 | 2.63 | 3.58 | 201 | 3.16 | 2.56 | 3.91 | 113 | 2.95 | 2.42 | 3.60 | 112 | 2.74 |
| BMI (kg/m<sup>2</sup>) | 35 vs 25 | 1.07 | 0.89 | 1.28 | 1 | 1.09 | 0.90 | 1.34 | 1 |  |  |  |  |  |
| HR (bpm) | 103 vs 78 | 1.16 | 1.05 | 1.29 | 8 | 1.19 | 0.97 | 1.45 | 3 |  |  |  |  |  |
| SBP (mmHg) | 150 vs 118 | 0.76 | 0.65 | 0.89 | 12 | 0.86 | 0.73 | 1.01 | 3 |  |  |  |  |  |
| RR (/min) | 23 vs 16 | 1.98 | 1.77 | 2.21 | 150 | 1.63 | 1.34 | 1.99 | 24 | 1.91 | 1.63 | 2.23 | 64 | 1.82 |
| Saturation (%) | 97.5 vs 94 | 0.61 | 0.52 | 0.72 | 37 | 0.77 | 0.65 | 0.90 | 10 |  |  |  |  |  |
| Temperature (°C) | 38 vs 37 | 1.16 | 1.02 | 1.32 | 5 | 1.05 | 0.87 | 1.27 | 0 |  |  |  |  |  |
| CRP (mg/L) | 118 vs 10 | 2.76 | 2.35 | 3.25 | 149 | 1.54 | 1.22 | 1.93 | 14 | 1.57 | 1.27 | 1.93 | 18 | 1.52 |
| LDH (U/L) | 322 vs 200 | 2.17 | 1.99 | 2.36 | 309 | 1.83 | 1.62 | 2.06 | 99 | 1.85 | 1.66 | 2.05 | 127 | 1.77 |
| Leucocytes (x10<sup>9</sup>/L) | 12.7 vs 6.7 | 1.01 | 0.92 | 1.11 | 0 | 0.88 | 0.75 | 1.02 | 3 |  |  |  |  |  |
| Lymphocytes (x10<sup>9</sup>/L) | 1.6 vs 0.66 | 0.67 | 0.60 | 0.75 | 48 | 1.03 | 0.90 | 1.19 | 0 |  |  |  |  |  |
| Albumin (g/L) | 42 vs 36 | 0.58 | 0.53 | 0.62 | 191 | 0.80 | 0.71 | 0.90 | 14 | 0.77 | 0.69 | 0.86 | 21 | 0.78 |
| Bicarbonate (mmol/L) | 25.9 vs 21.4 | 0.71 | 0.65 | 0.78 | 54 | 0.81 | 0.71 | 0.92 | 10 |  |  |  |  |  |
| Creatinine (μmol/L) | 111 vs 66 | 1.58 | 1.46 | 1.71 | 133 | 0.85 | 0.72 | 1.00 | 4 |  |  |  |  |  |
| Eosinophils (x10<sup>9</sup>/L) | 0.1 vs 0.004 | 0.77 | 0.64 | 0.93 | 7 | 1.08 | 0.90 | 1.30 | 1 |  |  |  |  |  |
| MCV (fL) | 94 vs 87 | 1.18 | 1.08 | 1.29 | 14 | 1.10 | 0.99 | 1.23 | 3 |  |  |  |  |  |
| Monocytes (x10<sup>9</sup>/L) | 0.95 vs 0.44 | 0.84 | 0.74 | 0.96 | 7 | 1.08 | 0.87 | 1.35 | 1 |  |  |  |  |  |
| Neutrophils (x10<sup>9</sup>/L) | 9 vs 2.2 | 0.97 | 0.88 | 1.07 | 0 | 0.94 | 0.82 | 1.08 | 1 |  |  |  |  |  |
| Sodium (mmol/L) | 140 vs 135 | 0.98 | 0.96 | 1.01 | 1 | 1.03 | 0.98 | 1.09 | 1 |  |  |  |  |  |
| Urea (mmol/L) | 9.7 vs 4.6 | 2.48 | 2.24 | 2.76 | 291 | 1.79 | 1.43 | 2.24 | 26 | 1.61 | 1.42 | 1.83 | 53 | 1.56 |

Consequently, to avoid overestimation of the discriminative ability, we limited validation of models in the development data to patients who were admitted from April 2020 onward.

D-dimer was not analyzed in the regression analysis, because 64% and 76% were missing in the development and validation data, respectively (Table 1). Based on a full model with restricted cubic splines of all potential variables, we decided to transform all biomarkers and RR with the natural logarithm, while keeping all other predictor effects linear. Some strong univariable associations with death – for example of lymphocytes and creatinine (Table 2; Wald statistics 48 and 133, respectively) – were very weak in multivariable analysis (Table 2; Wald statistics 0 and 4, respectively). The predictive ability of the resulting full multivariable regression model was mainly driven by age, LDH, urea, RR, CRP, Albumin, oxygen saturation and bicarbonate (ORs and Wald statistics in Table 2). A simple model – named COVID Outcome Prediction in the Emergency Department (COPE) – with linear age and logarithmic transforms of RR, CRP, LDH, albumin and urea captured most of the ability to predict death within 28 days (Table 2). Based on internal bootstrap validation we applied a shrinkage factor of 0.93 to the regression coefficients.

COPE showed good discrimination for predicting death in 4,498 patients who were admitted from April up to and including August 2020 in the first wave (Supplementary Figure 1; AUC in 4 hospitals 0.85 [95% confidence interval: 0.81; 0.88]; 0.81 [0.71; 0.91]; 0.86 [0.82; 0.90]; 0.85 [0.81; 0.88]) and, more importantly, in the validation sample of 3,235 patients who were admitted in the second wave from September up to and including December 2020 (Figure 2; AUC in 4 hospitals: 0.82 [0.78; 0.86]; 0.82 [0.74; 0.90]; 0.79 [0.70; 0.88]; 0.83 [0.79; 0.86]). The decrease in AUC over time was partly driven by less case mix heterogeneity – expressed by a lower model-based AUC (mbc) – of second wave patients (Figure 1; mbc in 4 hospitals: 0.81; 0.82; 0.81; 0.82) as compared to first wave patients (Supplementary Figure 1; mbc in 4 hospitals 0.82; 0.85, 0.83, 0.84). COPE was well-calibrated in second wave patients of each of the 4 hospitals, both on average – expressed by hospital-specific calibration intercepts: 0.08 [-0.15; 0.30]; -0.17 [-0.65; 0.30]; -0.01 [-0.40; 0.39]; -0.12 [-0.30; 0.07] – and by predicted risk levels – expressed by hospital-specific calibration slopes: 1.09 [0.86; 1.31]; 0.90 [0.49; 1.32]; 0.91 [0.57; 1.25]; 0.97 [0.79; 1.14] (Figure 2).

**Figure 1.**
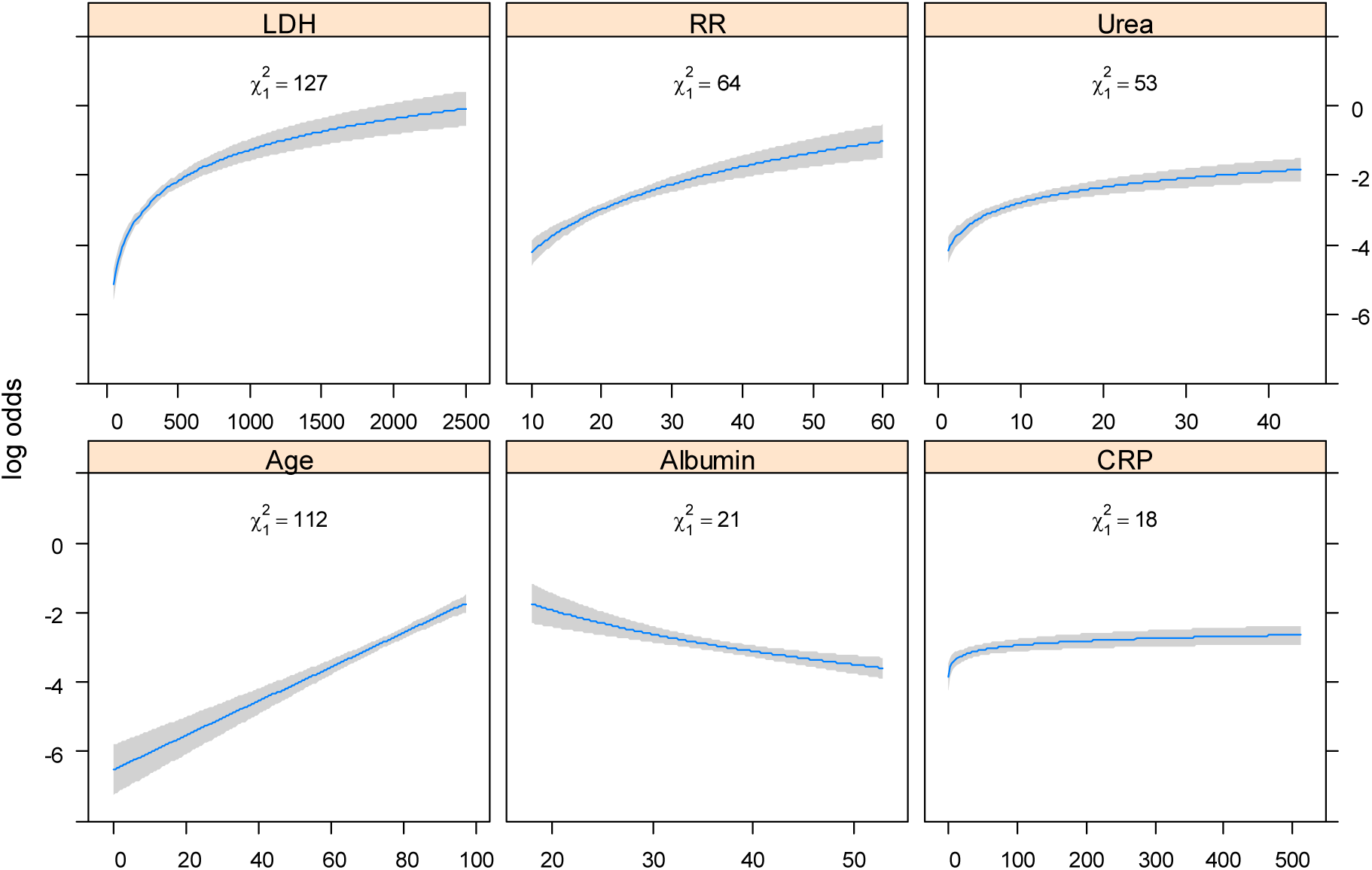
Multivariable effects of continuous predictors of death within 28 days. Predictions of the logarithm of the odds by continuous predictor levels, with other predictor levels set to the median. Wald statistics are listed within each plot to express variable importance.

**Figure 2.**
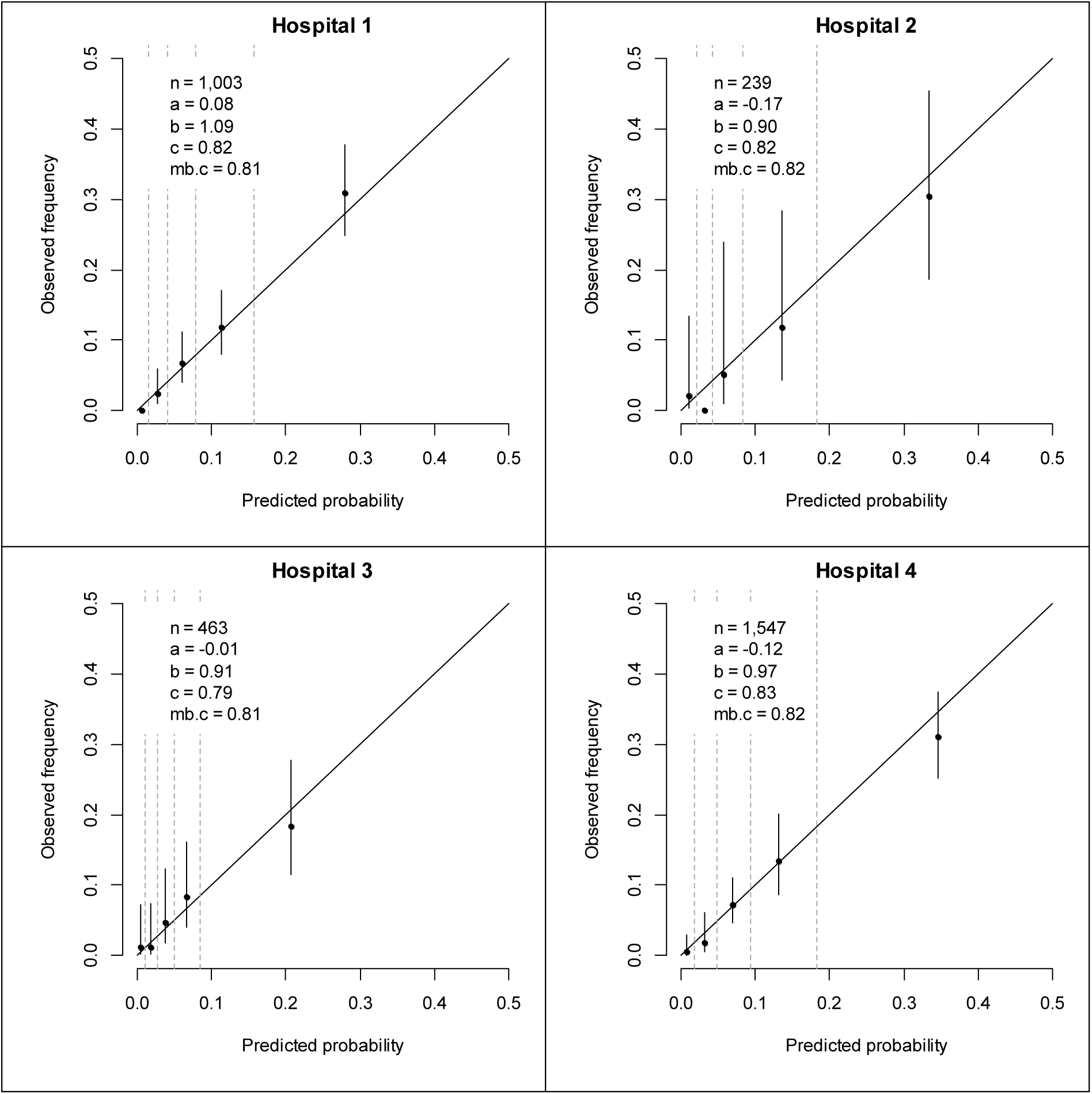
Temporal validation: Performance of COPE for predicting death in second wave patients. Calibration plots of patients who were admitted since September 2020 in 4 separate Dutch hospitals. n is number of patients; a = calibration intercept; b = calibration slope; c = AUC; mb.c = model-based AUC.

### Prediction of ICU admission

The probability of being admitted to the ICU was decreasing with age after the age of 70, likely reflecting the decision not to admit older patients to the ICU. When adjusting for this decreasing age effect after the age of 70 – by including a linear spline with a knot at age 70 in the regression model (Supplementary figure 2) – the strongest predictors of death were also predictive of ICU admission within 28 days, but associations were generally weaker for the latter (Table 3 vs Table 2). In patients below the age of 70, admitted from April up to and including August 2020, a model with the linear predictor of death calibrated to ICU admission had similar discriminative ability to a model that refitted all the predictor effects (AUC 0.71 for both models). For robustness, we implemented the calibrated model and not the refitted model (calibration slope 0.60; 95% confidence interval: 0.49; 0.70) into COPE for predicting ICU admission. To predict the need for ICU admission – rather than historically observed ICU admission – COPE ignores the decreasing age effect after the age of 70 when making predictions for future patients, since the observed ICU admission rate is probably an inaccurate estimate of the medical need for ICU admission. Thus, predictions of ICU admission after the age of 70 are based on an extrapolation of the observed age effect on ICU admission in patients below the age of 70. Due to the weaker predictor effects, the discriminative ability of COPE was more moderate for predicting ICU admission than for predicting death (Supplementary figure 3; AUC in 2 hospitals: 0.66 [0.58; 0.74]; 0.79 [0.69;0.88]). Although COPE significantly overestimated ICU admission in second wave patients (Figure 3; calibration intercept in 2 hospitals: -0.41 [-0.77; -0.05]; -0.72 [-1.34; -0.11]), it was better able to identify the patients at high risk of needing ICU admission, as expressed by higher discriminative ability (Figure 3; AUC in 2 hospitals: 0.84 [0.78; 0.90]; 0.81 [0.66; 0.95]) and substantially stronger predictor effects (calibration slope in 2 hospitals: 1.55 [1.03; 2.06]; 1.53 [0.60; 2.46]).

**Table 3.** Multivariable associations between predictors and ICU admission within 28 days. Odds ratios (OR) with 95% confidence intervals (CI) for separate variables (columns “Univariable”) and for a model with the six strongest predictors of death, corrected for a decreasing probability of ICU admission after the age of 70 (columns “Multivariable”). Variable importance is expressed with the Wald statistic (columns “Wald”). Associations are based on 2,633 patients of whom 214 were admitted to the ICU within 28 days.

| Predictor | Contrast | Univariable |  |  |  | Multivariable |  |  |  |
| --- | --- | --- | --- | --- | --- | --- | --- | --- | --- |
|  |  | OR | 95% CI |  | Wald | OR | 95% CI |  | Wald |
| Month | ≥April vs <April | 2.06 | 1.51 | 2.81 | 21 | 1.63 | 1.16 | 2.28 | 8 |
| Age (years) | 80 vs 58 | 1.96 | 1.47 | 2.62 | 21 | 1.76 | 1.32 | 2.35 | 15 |
| RR (/min) | 23 vs 16 | 1.76 | 1.48 | 2.09 | 40 | 1.71 | 1.40 | 2.09 | 27 |
| CRP (mg/L) | 118 vs 10 | 1.88 | 1.44 | 2.44 | 22 | 1.30 | 0.95 | 1.77 | 3 |
| LDH (U/L) | 322 vs 200 | 1.73 | 1.52 | 1.98 | 66 | 1.44 | 1.25 | 1.67 | 24 |
| Albumin (g/L) | 42 vs 36 | 0.75 | 0.64 | 0.88 | 13 | 0.95 | 0.78 | 1.17 | 0 |
| Urea (mmol/L) | 9.7 vs 4.6 | 1.29 | 1.08 | 1.54 | 8 | 1.36 | 1.10 | 1.66 | 8 |
| **Adjusted for:** |  |  |  |  |  |  |  |  |  |
| Max[Age-70, 0] (years) * | 80 vs 58 | 0.20 | 0.13 | 0.30 | 57 | 0.17 | 0.11 | 0.27 | 65 |

\* In univariable regression, only the association of age with ICU admission was adjusted for Max[Age-70, 0]

**Figure 3.**
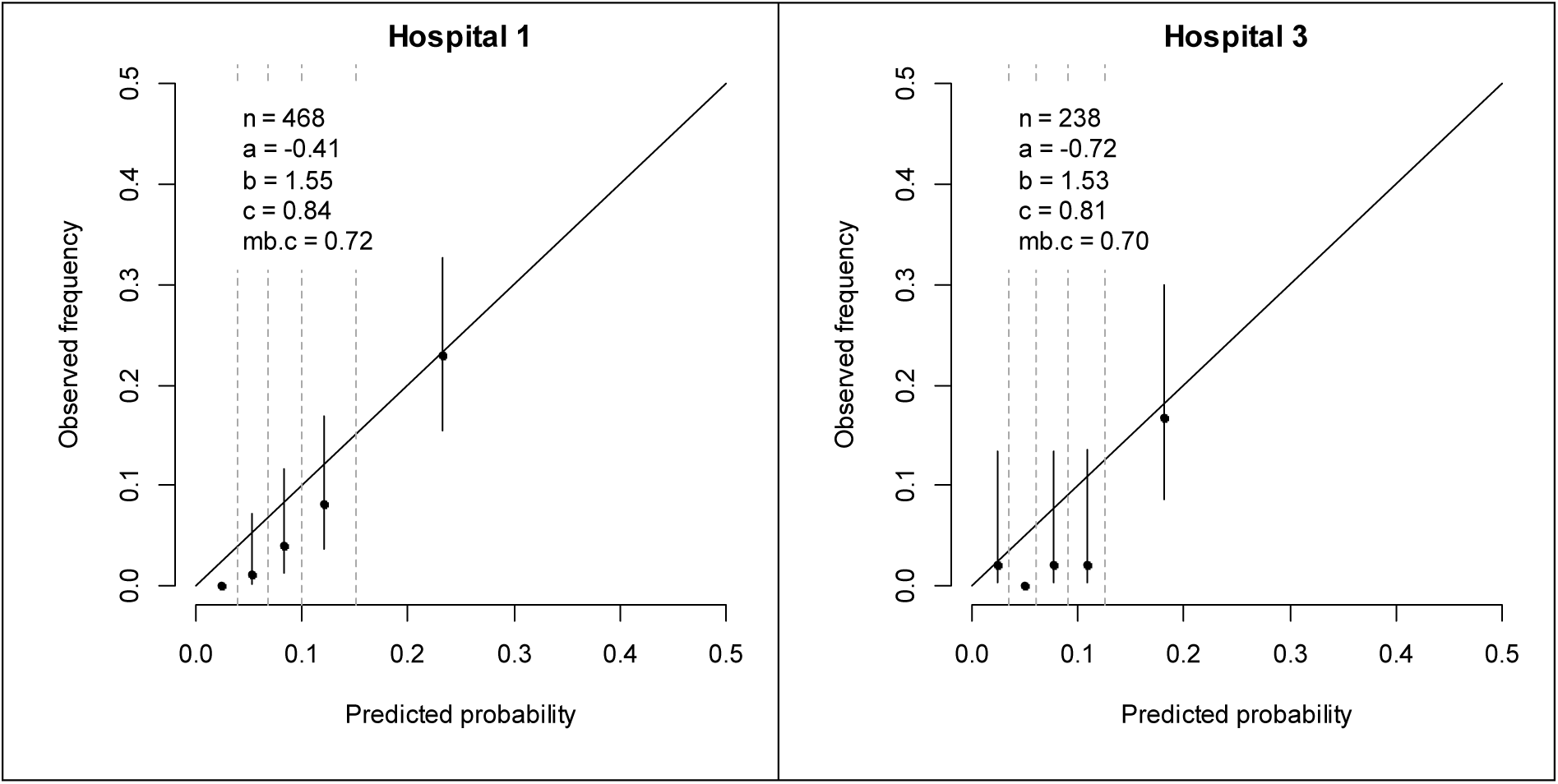
Temporal validation: Performance of COPE for predicting ICU admission in second wave patients. Calibration plots of patients who were admitted since September 2020 in 2 separate Dutch hospitals. n is number of patients; a = calibration intercept; b = calibration slope; c = AUC; mb.c = model-based AUC.

### Model presentation

The resulting COPE model for predicting death as well as need for ICU admission within 28 days after hospital admission (formulas in Figure 4) are implemented as a publicly accessible web-based application (https://mdmerasmusmc.shinyapps.io/COPE/) and as independent mobile apps (“COPE Decision Support”). For optimal transparency, the web and mobile applications include a detailed description of the derivation of COPE (Supplement 1), descriptions of the data that were used for development and validation of COPE, and calibration plots of temporal validation in the separate hospitals.

**Figure 4.**
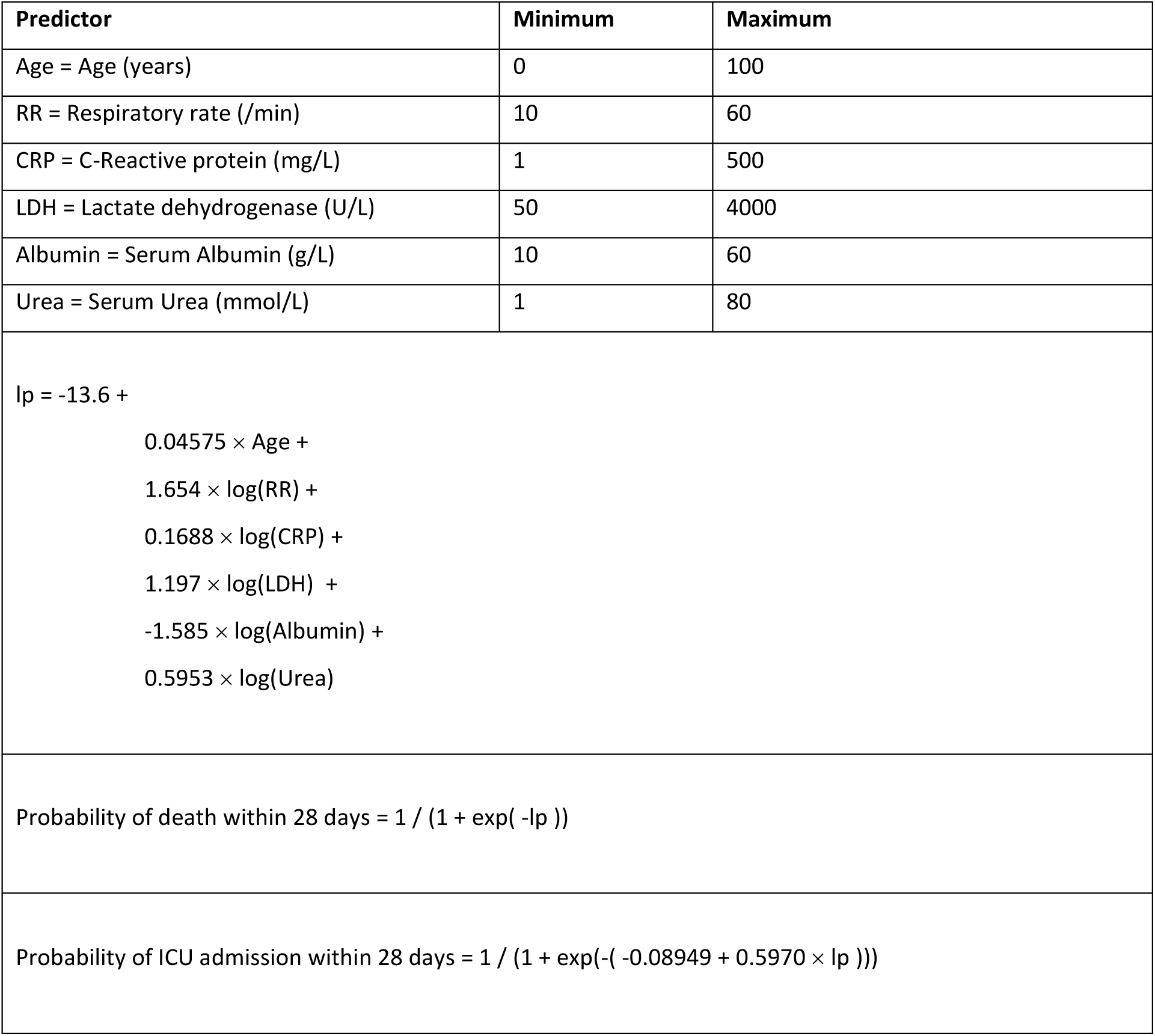
COPE definition. Implemented in web application https://mdmerasmusmc.shinyapps.io/COPE/ and mobile application “COPE Decision Support”. log = natural logarithm; exp = natural exponential

According to the Transparent Reporting of a multivariable prediction model for Individual Prognosis Or Diagnosis (TRIPOD) checklist (Supplementary table 1), all relevant items are covered in this manuscript, except for the availability of data sets (14, 15). Data transfer agreements with each of the contributing hospitals preclude any sharing of data sets.

## DISCUSSION

We developed COPE for prediction of in-hospital death and need for intensive care when patients with suspected COVID-19 present at the Emergency Department. Developed in patient data from the first wave of the pandemic, based on six quickly and objectively obtainable predictors – age, respiratory rate, LDH, CRP, albumin and urea – COPE discriminated well and was well-calibrated in patients admitted to hospitals in the second wave of the pandemic, both for predicting in-hospital death and for ICU admission.

The clinical presentation of COVID-19 is broad and varies from asymptomatic to critical disease. Some patients who initially have mild symptoms progress to severe disease within one week (16). In the ED physicians need to identify high-risk patients – i.e. those at high risk of deterioration and/or death – requiring treatment in the ICU, intermediate-risk patients requiring admission to the clinical ward, and low-risk patients who can potentially be sent home. Since COPE is based on data that are routinely measured, or at least readily available in the ED, it can act as a tool to support such decisions. Hospitalized patients who are at high risk for mortality or need for ICU admission should be more intensively watched, and when a high load of high-risk patients occurs in the ED, this should be taken into account in the ICU capacity planning.

We requested 19 large Dutch hospitals to supply retrospective data on the cohorts of COVID-19 patients who were admitted to their hospital. This request for data was sent out very early in the pandemic and was greeted with enthusiasm. Probably due to the enormous pressure on health care at that time, 4 hospitals supplied useable data for the analysis. The contributing hospitals were well-spread over the Netherlands, with one in the west, two in the south, and one in the east of the country, and are a mix of academic and large teaching hospitals. we believe they are representative for health care in the Netherlands. Although the consistently good performance of COPE across the hospitals may support its generalizability to other countries, geographic validation would be additionally reassuring, since the epidemic, and clinical practice for this novel disease, may have substantial inter-country variability.

COPE was developed based on 5,831 patients of whom 629 died within 28 days. This effective sample size of 629 events was ample to start the development process with a full model of 45 regression coefficients (14 events per variable), that is one binary predictor (sex) and 22 continuous predictors with 2 regression coefficients – due to using non-linear terms – each (17). To prevent too extreme predictions of COPE in new data, we applied a shrinkage factor to its regression coefficients, based on a bootstrap procedure with backward selection starting from the full model (11).

Our explicit aim was to develop a score based on quickly and objectively obtainable predictors at presentation at the ED. Consequently, pre-existing comorbidities, the level of consciousness measured by the Glasgow Coma Scale, and chest radiography results – although predictive for outcomes of COVID patients in other studies – were not considered here (4, 6, 7). Some predictors were promising in univariable analysis, such as lymphocytes and creatinine, but had negligible effects in multivariable analysis, because of strong correlations with other, more important predictors. Other predictors, such as oxygen saturation and bicarbonate, were significantly associated with death in multivariable analysis, but were not selected into the final model, since our explicit aim was to develop a simple model and the incremental value of these predictors was minimal. To achieve this aim, we only selected the strongest predictors – age, respiratory rate, LDH, CRP, albumin and urea – resulting in a parsimonious but well-performing model.

Besides mortality, we aimed to predict the need for ICU admission. A limitation of our study is that the *need for* ICU admission differs from the *observed decisions on* ICU admission, and is inherently difficult to model, because recorded ICU admissions express historical decisions at national, regional, hospital or even intensivist level. As a robust solution, we exploited the strong correlation between need for intensive care and death, by calibrating our model for predicting death to the observed ICU admissions, adjusting for a linear decrease with age after the age of 70. Hence, we assumed a linear relationship between (the logarithm of the odds of) death and need for ICU admission, and that all patients below the age of 70 needing intensive care were actually admitted to the ICU, that is *the need for ICU admission* is well estimated by the *observed decisions on ICU admission* for patients below the age of 70. The latter is reasonable given the sufficiency of ICU beds for Dutch patients throughout the pandemic. The discriminative ability of this re-calibration approach was very similar to that of a model that refitted all associations between COPE predictors and ICU admission. With temporal validation in 2 separate hospitals, we showed that COPE discriminated very well between patients at low and high risk of ICU admission and that the predicted probability of ICU admission was well-calibrated for the 20% highest-risk patients. Nevertheless, recalibration of COPE for predicting need for ICU admission to local circumstances may be necessary.

The absence of external validation in our study – measuring the predictive performance of COPE in hospitals that were not present in the development data – may be considered a limitation of this study (18). However, the combination of temporal validation – in second wave patients – and geographic validation – in separate hospitals – is a strength of this study (19). Although COPE already performed very well when validated across time and space, future research should focus on analyses of potential time trends not captured by the predictors – e.g. decreases in mortality thanks to improvements in treating COVID-19 patients (20) –, potential changes in predictor effects in time (interactions between predictors and time), and the impact of potential differences in patient case mix in countries other than the Netherlands (international validation). These case mix differences should primarily affect calibration, requiring an update of the model intercept, but not discrimination.

In conclusion, COPE, a simple model based on 6 quickly and objectively obtainable predictors in the ED, is well able to predict mortality and need for ICU admission for patients who present to the ED with suspected COVID-19. COPE may support patients and doctors in decision making.

## Data Availability

According to data transfer agreements the data is not publicly available.

## ACKNOWLEDGMENTS

We thank Noreen van der Linden and the Dutch Network of Acute Care (LNAZ) for support with collecting the data.

## AUTHORS’ CONTRIBUTIONS

David van Klaveren, Hester Lingsma, Jelmer Alsma, Rob JCG Verdonschot, Hilde RH de Geus, Rozemarijn L Van Bruchem-Visser, Jelle R Miedema, Annelies Verbon, Els van Nood, David M Kent and Stephanie CE Schuit conceived and designed the study. Jelmer Alsma, Rob JCG Verdonschot, Dick TJJ Koning, Marlijn JA Kamps, Tom Dormans, Robert Stassen, Sebastiaan Weijer, Klaas-Sierk Arnold and Benjamin Tomlow were responsible for collecting the data. David van Klaveren analyzed the data and wrote the first draft of the paper. Alexandros Rekkas implemented the models into a web application. All authors contributed to writing the paper and approved the final version.

The corresponding author attests that all listed authors meet authorship criteria and that no others meeting the criteria have been omitted. The Corresponding Author has the right to grant on behalf of all authors and does grant on behalf of all authors, a worldwide licence to the Publishers and its licensees in perpetuity, in all forms, formats and media (whether known now or created in the future), to i) publish, reproduce, distribute, display and store the Contribution, ii) translate the Contribution into other languages, create adaptations, reprints, include within collections and create summaries, extracts and/or, abstracts of the Contribution, iii) create any other derivative work(s) based on the Contribution, iv) to exploit all subsidiary rights in the Contribution, v) the inclusion of electronic links from the Contribution to third party material where-ever it may be located; and, vi) licence any third party to do any or all of the above.”

## ETHICS APPROVAL

The Daily Board of the Medical Ethics Committee Erasmus MC of Rotterdam, The Netherlands, has approved the research proposal (MEC-2020-0297).

## TRANSPARENCY STATEMENT

The lead author (David van Klaveren) affirms that the manuscript is an honest, accurate, and transparent account of the study being reported; that no important aspects of the study have been omitted; and that any discrepancies from the study as originally planned have been explained.

All authors have completed the ICMJE uniform disclosure form at www.icmje.org/coi_disclosure.pdf and declare: no support from any organisation for the submitted work; no financial relationships with any organisations that might have an interest in the submitted work in the previous three years; no other relationships or activities that could appear to have influenced the submitted work.

---

**Supplementary table 1.**
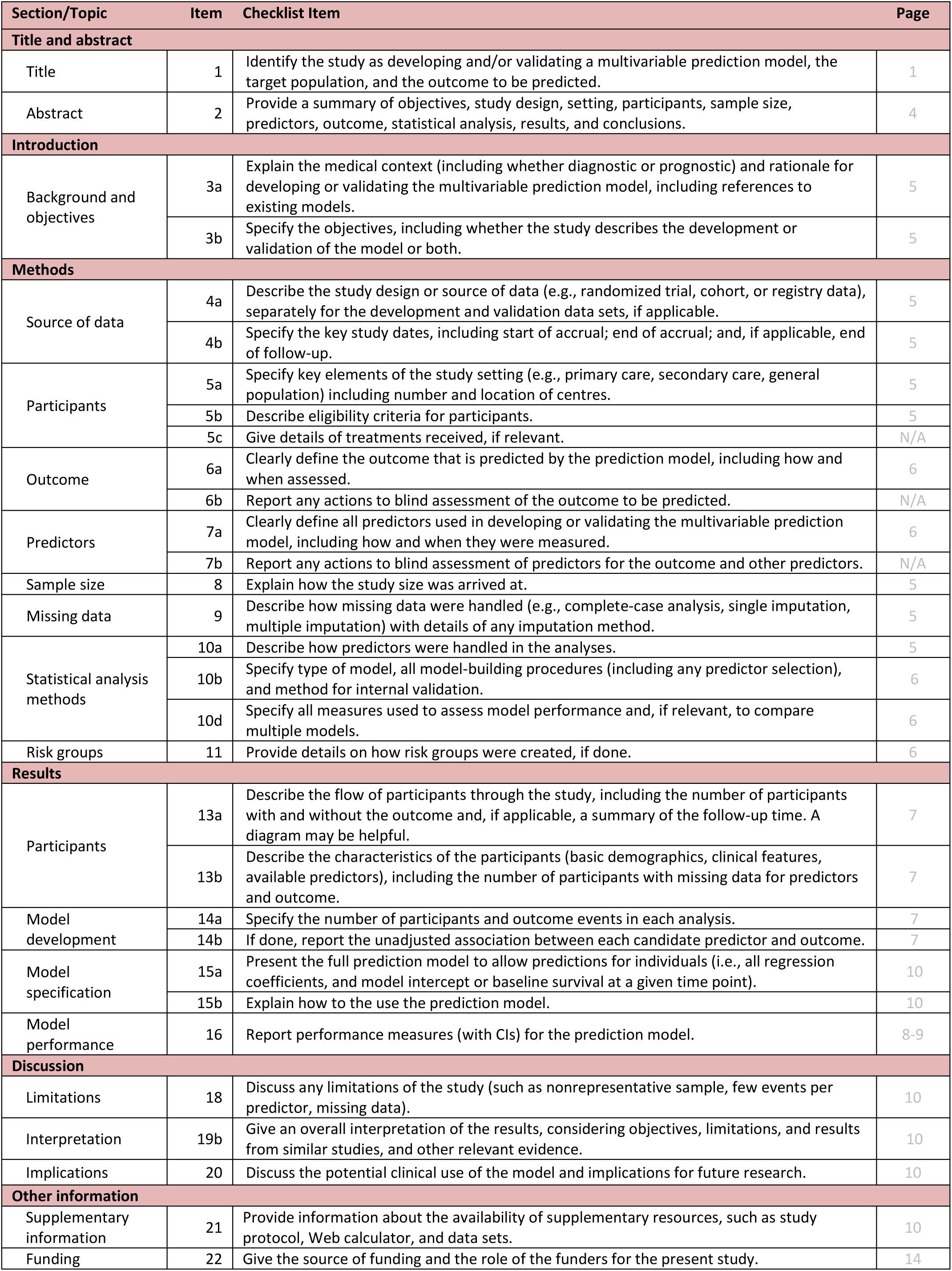
TRIPOD Checklist Prediction Model Development.

**Supplementary figure 1.**
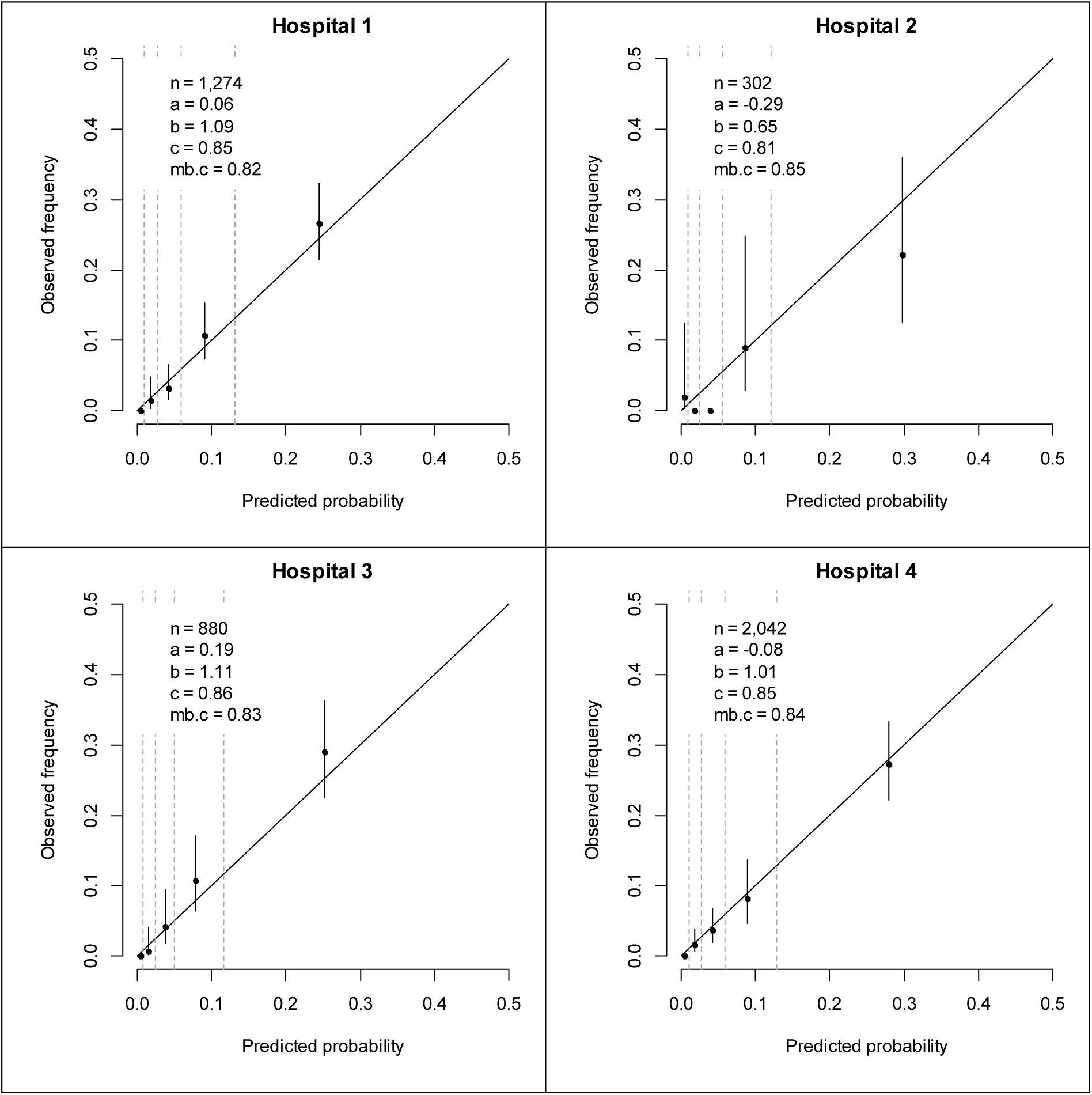
Apparent validation: Performance of COPE for predicting death in first wave patients. Calibration plots of patients who were admitted from April up to and including August 2020 in 4 separate Dutch hospitals. n is number of patients; a = calibration intercept; b = calibration slope; c = AUC; mb.c

**Supplementary figure 2.**
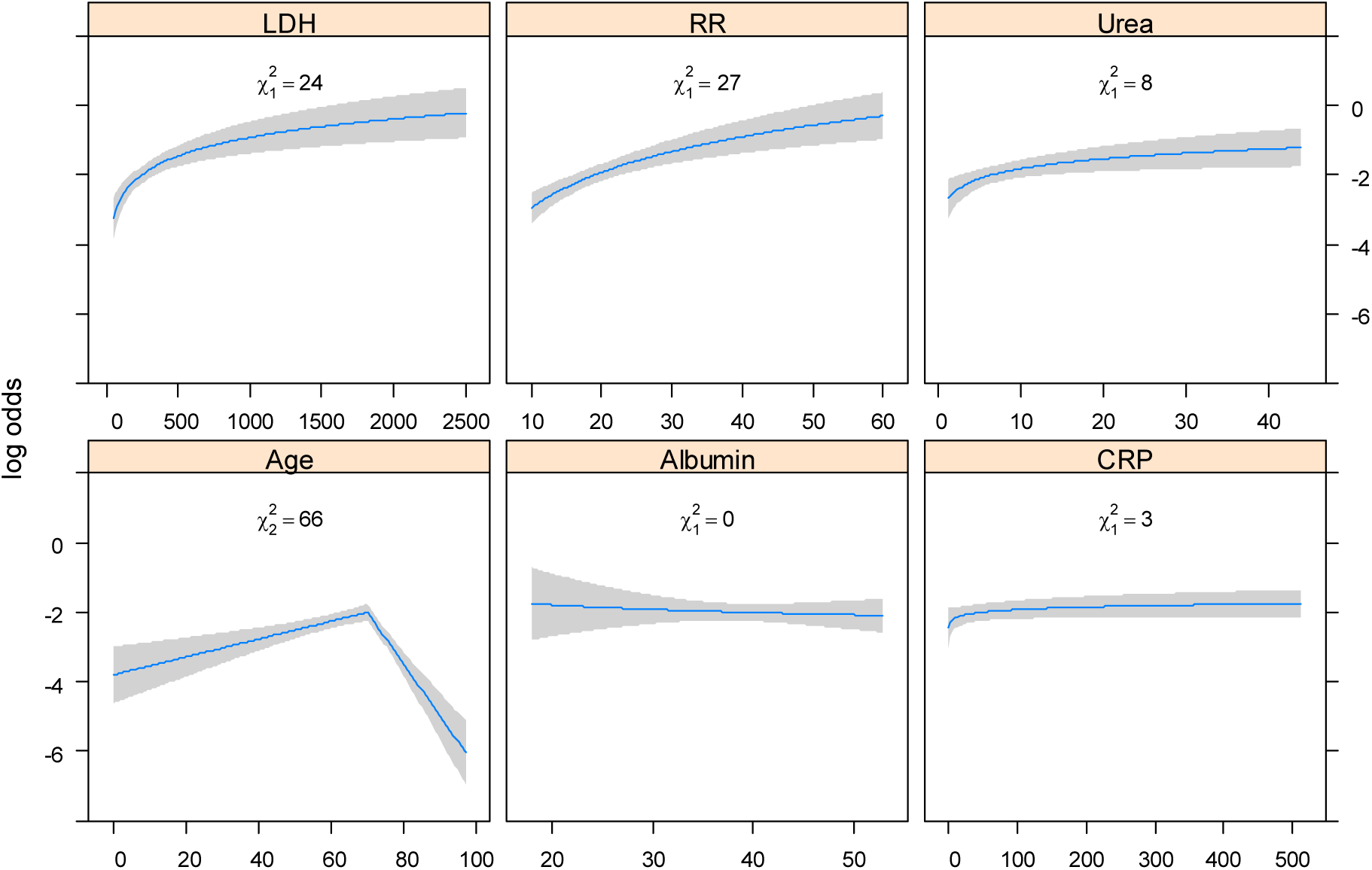
Multivariable effects of continuous predictors of ICU admission within 28 days. Predictions of the logarithm of the odds by continuous predictor levels, with other predictor levels set to the median. Age is modelled with a linear spline with a knot at age 70. Wald statistics are listed within each plot to express variable importance.

**Supplementary figure 3.**
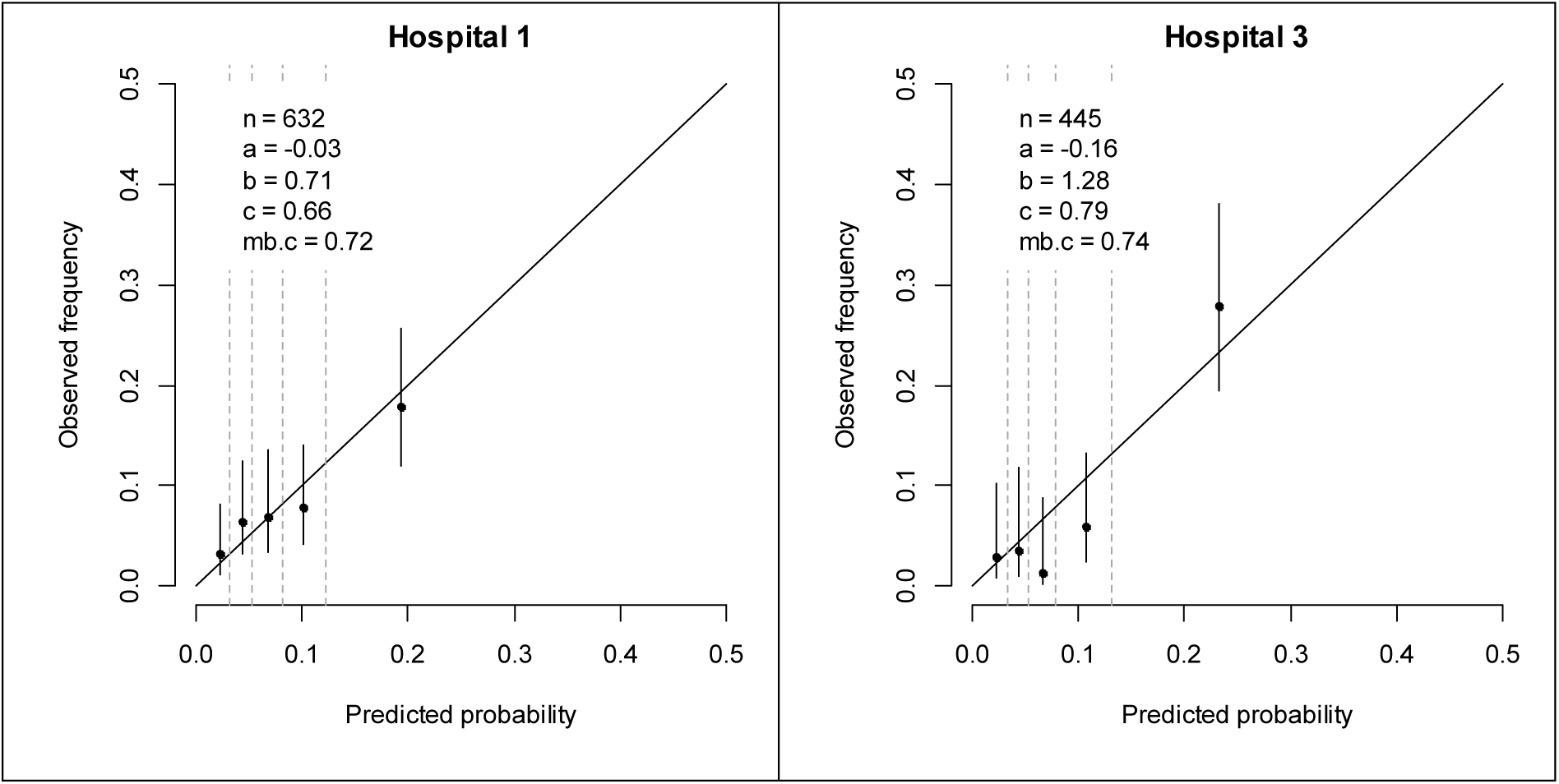
Apparent validation: Performance of COPE for predicting ICU in first wave. Calibration plots of patients who were admitted from April up to and including August 2020 in 2 separate Dutch hospitals. n is number of patients; a = calibration intercept; b = calibration slope; c = AUC; mb.c = model-based AUC.

## Supplement 1 Description COPE web application

### Background and aim

The COVID-19 pandemic is putting extraordinary pressure on emergency departments (EDs). Clinical prediction models have the potential to support decision making about hospital admission, but currently available models were recently assessed to contain a high risk of bias. We aimed to develop a simple and valid model for predicting mortality and need for ICU in patients who are suspected to have COVID-19 when presenting at the ED.

### Methods

For model development, we included patients that presented at the ED and were admitted to 4 large Dutch hospitals with suspected COVID-19 between March and August 2020, the first wave of the pandemic in the Netherlands. Patients being transferred from or to other hospitals were excluded since information on predictors or outcomes was missing. The outcomes of interest were death and admission to ICU within 28 days. Based on prior literature we included patient characteristics (sex, age, BMI), vital parameters (oxygen saturation, systolic blood pressure, heart rate, respiratory rate [RR], body temperature) and blood test values (C-reactive protein [CRP], lactic dehydrogenase [LDH], D-Dimer, leucocytes, lymphocytes, monocytes, neutrophils, eosinophils, MCV, albumin, bicarbonate, creatinine, sodium, urea), all measured at ED admission, as potential predictors. Further we included month of admission to capture changes in outcomes over time. Logistic regression was used to obtain predicted probabilities of death and of being admitted to the ICU, both within 28 days after admission. Model performance was assessed with temporal validation in patients who presented between September and December 2020 (second wave). We assessed discriminative ability with the area under the operator receiver characteristic curve (AUC) and calibration with calibration plots, calibration intercepts, and calibration slopes. We used multiple imputation to account for missing predictor values.

### Results

The development data included 5,831 patients who presented and were admitted at the ED up until August 2020, of whom 629 (10.8%) died and 5,070 (86.9%) were discharged within 28 days after admission. A simple model – named COVID Outcome Prediction in the Emergency Department (COPE) – with linear age and logarithmic transforms of RR, CRP, LDH, Albumin and Urea captured most of the ability to predict death within 28 days. Patients who were admitted in the first month of the pandemic in the Netherlands had substantially increased risk of death (odds ratio 2.06; 95% confidence interval 1.68-2.52). COPE was well-calibrated and showed good discrimination for predicting death in 3,252 patients in the second wave (AUC in 4 hospitals: 0.82; 0.82; 0.79; 0.83). Admission to ICU was fully recorded for 2,633 first wave patients in 2 hospitals (214 ICU admissions within 28 days). The same predictors captured most of the ability to predict ICU admission within 28 days. However, after the age of 70, the probability of being admitted to the ICU was decreasing with age, probably reflecting the decision not to admit older patients to the ICU. To predict the need for ICU admission – rather than historically observed ICU admission – we kept a linear (decreasing) age effect after the age of 70 in the model, which will be ignored when making future predictions. COPE was well able to identify patients at high risk of needing IC in second wave patients below the age of 70 (AUC 0.84; 0.81), but overestimated ICU admission for low-risk patients. The models are implemented as a web-based application.

### Conclusion

COPE, a simple tool based on 6 routinely measured predictors in the ED, is well able to predict mortality and ICU admission for patients who present to the ED with suspected COVID-19. COPE may help to inform patients and doctors when deciding on hospital admission.

